# Multimodal brain age prediction reveals dissociable signatures of health, cognition and disease risk in 24,648 UK Biobank participants

**DOI:** 10.64898/2026.06.29.26355335

**Authors:** Ruize Yu, Shanshan Shao, Feng Xu

## Abstract

Deep-learning brain-age models usually rely on a single MRI contrast, although brain aging affects grey matter, white matter, iron-rich nuclei and cerebrovascular tissue. Here we trained 3D DenseNet121 models to predict chronological age from five MRI modalities - T1, T2 FLAIR, T1+T2 early fusion, diffusion MRI and susceptibility-weighted imaging - in up to 24,648 UK Biobank participants, with external validation in the Parkinson’s Progression Markers Initiative. T1+T2 fusion achieved the lowest error (mean absolute error 2.19 yr; Pearson r = 0.934), but downstream analyses showed modality-specific signals. Diffusion MRI brain age gap was uniquely associated with arterial stiffness and most strongly predicted incident type 2 diabetes; susceptibility-weighted imaging showed the largest cognitive effect sizes; T2 FLAIR best predicted all-cause dementia and cerebrovascular disease; and diffusion MRI carried the highest hazard for Alzheimer’s disease. Grad-CAM indicated that fusion shifted attribution from grey to white matter, and Mendelian randomization implicated physical activity, diet, smoking and alcohol intake as causal lifestyle determinants. These findings establish brain age as a family of modality-specific biomarkers.

## Introduction

Global life expectancy has risen markedly over the past century, but the additional years are increasingly burdened by age-related neurological and psychiatric morbidity ^1^. Brain aging is the central biological process underlying this burden, yet trajectories vary markedly between individuals of identical chronological age, making chronological age an imperfect proxy for neurobiological health ^2;3^. This dissociation has driven a search for individualised biomarkers that capture the actual pace of neural senescence ^2;4;5^.

The brain-age paradigm — in which a model is trained to predict chronological age from neuroimaging data and the residual (predicted minus chronological age, hereafter brain age gap, BAG) is used as an individual-level biomarker — has become one of the most widely studied imaging-derived phenotypes in computational neuroscience ^6–8^. Early implementations used classical machine learning on pre-segmented structural features and achieved MAE values of 4–6 yr ^7^, while modern three-dimensional convolutional networks — particularly DenseNet, whose dense connectivity mitigates vanishing gradients in volumetric inputs — learn predictive features directly from minimally processed brain images ^9–12^. Peng et al. ^10^ showed that a lightweight fully convolutional network trained on 11,675 UK Biobank T1 scans reached an MAE of 2.14 yr; subsequent DenseNet- and ResNet-based models have converged on MAE values of 2.0–2.5 yr on T1-weighted UK Biobank data, defining the current state of the art^13–15^.

Three fundamental questions remain unresolved. First, nearly all deep-learning brain-age models are trained on a single MRI contrast — predominantly T1-weighted structural imaging — even though brain aging is a multi-compartment process involving grey matter atrophy, white matter microstructural degradation, iron accumulation in subcortical nuclei and cerebrovascular pathology ^16–18^. T2 FLAIR reveals white matter hyperintensities of presumed vascular origin; dMRI captures white matter tract integrity; SWI is sensitive to non-haem iron and microhaemorrhages ^16;19^. Each contrast samples a distinct biological compartment, but whether these signals translate into dissociable phenotypic, cognitive and prognostic profiles within a unified brain-age framework has not been systematically established ^13;20;21^. Second, brain-age studies have typically focused on a narrow set of cognitive measures or a single disease endpoint, and observational lifestyle–BAG associations remain vulnerable to residual confounding and reverse causation; genetically instrumented causal inference has only recently been applied to brain-age phenotypes ^22;23^. Third, the neuroanatomical basis of deep-learning age predictions remains poorly characterised, and most studies report a single attribution method without quantifying how this choice constrains the interpretation^24–26^.

The UK Biobank (UKB) imaging enhancement, with multimodal brain MRI in >60,000 participants embedded in a deeply phenotyped prospective cohort and linkage to hospital episode statistics and mortality registries, provides an unprecedented platform to address these questions at scale ^16;27^. Here we capitalise on this resource to (i) train and systematically compare DenseNet121 brain-age models across five MRI modalities (T1, T2 FLAIR, T1+T2 early fusion, dMRI and SWI) in up to 24,648 participants, with external validation in PPMI; (ii) characterise the phenotypic, cognitive and disease-outcome associations of modality-specific BAG across 49 cognitive tests, 11 health phenotypes and 26 disease endpoints; (iii) contrast Grad-CAM attribution maps across all five modalities to establish the neuroanatomical correlates of multimodal age prediction; and (iv) use two-sample Mendelian randomization (MR) with independent genetic instruments to test causal effects of seven lifestyle exposures on BAG. We show that brain age is not a unitary construct but a family of dissociable modality-specific signatures, each with a distinct profile of health associations, cognitive correlates, disease risk, neuroanatomical basis and causal lifestyle determinants.

## Results

### Modality-specific brain-age models in UK Biobank

Of approximately 500,000 UKB participants aged 40–69 yr at recruitment, the imaging enhancement (initiated 2014) has acquired brain, cardiac and body MRI from a subset on four identical Siemens Skyra 3T scanners under a harmonised protocol ^16;19^. Brain imaging comprised T1-weighted MPRAGE (1 mm isotropic), T2 FLAIR (1.05 × 1 × 1 mm), dMRI (2 mm isotropic, 100 directions) and SWI (0.8 mm isotropic) in a single ∼35-min session. After applying UKB central QC and requiring complete covariate and genetic-ethnicity data, our analytical samples were 24,114 (T1), 23,421 (T2 FLAIR), 22,885 (T1+T2 fusion), 21,067 (dMRI) and 20,575 (SWI) participants; 18,107 had data on all five modalities and were used for cross-modality comparative analyses including disease-risk models (Fig. 1a). Mean age at imaging was 64.2 yr (s.d. 7.6); 52.7% were female. Age-decile distributions were balanced across modalities, supporting reliable cross-modality comparisons.

**Figure 1:**
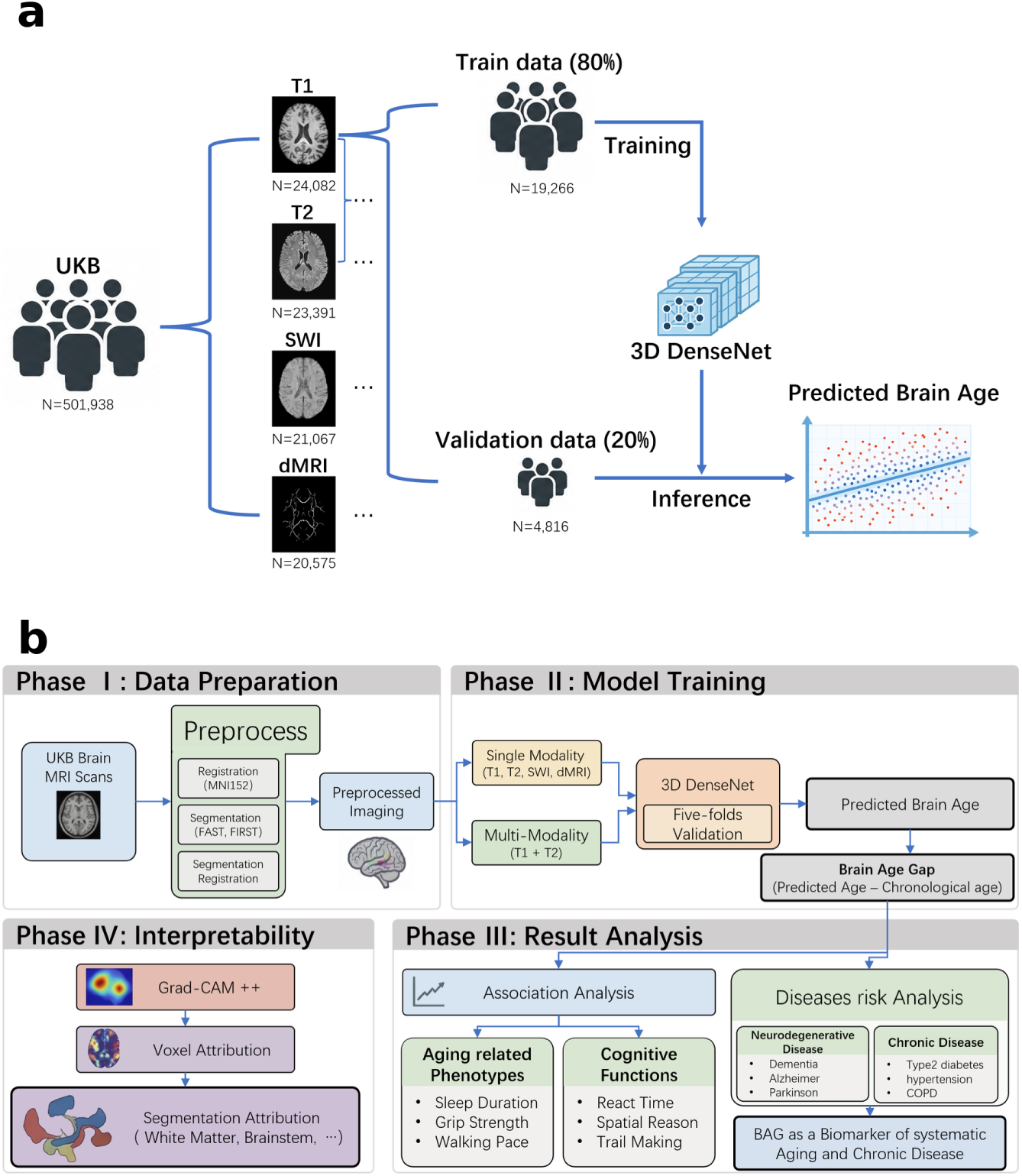
Study design and brain-age modelling workflow. **a**, Cohort assembly: UKB participants with each modality were split 80/20 into training and held-out validation sets within five-fold stratified cross-validation; final analytical samples for the five modalities are shown. **b**, Four-phase pipeline: pre-processing (MNI152 registration, FSL FAST/FIRST segmentation), DenseNet121 model training under single- or multi-modality input, Grad-CAM interpretability and downstream phenotypic/disease analyses. Images reproduced by kind permission of UK Biobank ©.

All five brain-age models used a 3D DenseNet121 backbone (MONAI/PyTorch) trained with five-fold cross-validation stratified by age decile, MAE loss, Adam optimiser (1 × 10^−4^ learning rate; cosine annealing), batch size 96 and distributed data parallel across three NVIDIA RTX 4090 GPUs (Fig. 1b; Methods). All inputs were registered to MNI152 1 mm^3^ stereotactic space using affine (12-parameter) transformation only.

Prediction performance varied substantially across modalities (Table 1). T1+T2 fusion, which concatenates the two image volumes at the input channel, achieved the lowest error (MAE 2.19 yr; Pearson r = 0.934; R^2^ = 0.873), followed by T2 FLAIR (MAE 2.27 yr; r = 0.929) and T1 (MAE 2.46 yr; r = 0.917). dMRI (MAE 2.81 yr; r = 0.892) and SWI (MAE 2.88 yr; r = 0.885) showed higher errors. The T1+T2 fusion MAE of 2.19 yr is lower than the 3D DenseNet of Bashyam et al. ^12^ and the multimodal models of Niu et al. ^20^ and Mouches et al. ^21^, and approaches the SFCN benchmark of Peng et al. ^10^ despite a larger and more heterogeneous sample.

**Table 1:** Brain-age prediction accuracy in UK Biobank and external validation in PPMI. UK Biobank metrics are from held-out folds of DenseNet121 models trained per modality with five-fold cross-validation. PPMI metrics are from applying the UKB-trained models to PPMI after identical affine-only registration to MNI152 1 mm. *Raw MAE* is the uncalibrated prediction error; *Cal. MAE* applies age-stratified isotonic calibration. External validation was performed for T1 and T2 FLAIR only; PPMI dMRI and SWI sequences are not protocol-compatible with UKB.

| Modality | UK Biobank (within-cohort) |  |  | PPMI (external) |  |  |  |
| --- | --- | --- | --- | --- | --- | --- | --- |
|  | *N* | MAE (yr) | *r* | *N* | Raw MAE | Cal. MAE | *r* |
| T1 | 24,114 | 2.46 | 0.917 | 3,151 | 6.19 | 4.75 | 0.689 |
| T2 FLAIR | 23,421 | 2.27 | 0.929 | 2,077 | 4.80 | 4.19 | 0.706 |
| T1+T2 fusion | 22,885 | 2.19 | 0.934 | — | — | — | — |
| dMRI | 21,067 | 2.81 | 0.892 | — | — | — | — |
| SWI | 20,575 | 2.88 | 0.885 | — | — | — | — |

The advantage of T2 FLAIR over T1 has not, to our knowledge, been systematically reported. T2 FLAIR is sensitive to white matter hyperintensities (WMHs), which accrue with age even in cognitively normal individuals and reflect small-vessel cerebrovascular pathology ^16;17^. The further improvement under T1+T2 fusion indicates that T1 and T2 carry partially non-redundant information about chronological age ^18;20^. BAG was bias-corrected per modality for regression-to-the-mean using the method of Liang et al. ^28^; after correction, BAG distributions were centred and showed s.d. 4.50–4.92 yr across modalities, with negligible residual correlation with chronological age (|r| < 0.02), ensuring downstream associations are not confounded by age-dependent prediction bias. Pairwise correlations among modalities ranged from r = 0.55 (dMRI vs. SWI) to r = 0.78 (T1 vs. T2), implying that ∼30–60% of BAG variance is shared while a substantial component is modality-specific — a quantitative argument for moving beyond single-modality BAG.

### External validation in PPMI

A critical preprocessing decision was the exclusive use of affine registration, without nonlinear (FNIRT) normalisation. In pilot experiments on the Parkinson’s Progression Markers Initiative (PPMI) cohort, FNIRT-based preprocessing reduced the Pearson correlation between predicted and chronological age to r = 0.40 — consistent with reports that aggressive spatial normalisation attenuates the biological signal available to predictive models ^29^. Switching to affine-only registration — matching the UKB pipeline ^19^ — restored r to 0.675 (T1) and 0.710 (T2 FLAIR) on PPMI, confirming that the models capture generalisable, non-registration-confounded age-related morphology rather than UKB-specific scanner or registration artefacts. External validation was confined to T1 and T2 FLAIR because the dMRI and SWI sequences in PPMI are not directly compatible with the UKB acquisition protocol ^16^.

### Phenotypic associations

We tested associations between modality-specific BAG and 11 health phenotypes plus 49 cognitive tests using ordinary least squares with robust (Huber–White) standard errors, adjusted for chronological age, sex, ethnicity, assessment centre and BMI; effect sizes are reported as standardised *β* with Benjamini–Hochberg false-discovery-rate (FDR) correction at q < 0.05.

Higher BAG was consistently associated with poorer physical health across modalities (Fig. 2). Grip strength — a well-established marker of biological aging and mortality risk ^4;18^ — showed robust negative associations with BAG in every modality (left hand *β* between −0.044 and −0.054; right hand *β* between −0.036 and −0.053; all P < 10^−4^). Self-rated poor health was positively associated with BAG in every modality (*β* between +0.016 and +0.030, all P < 0.02), with the strongest signal for dMRI. Self-reported slow walking pace was significantly associated with T2, dMRI and SWI BAG (*β* ≈ +0.020 to +0.025); basal metabolic rate (BMR) showed its largest negative association with T2 FLAIR BAG (*β* = −0.061, P = 3.4×10^−5^); and BMI was modestly positively associated with BAG (largest for dMRI, *β* = +0.024, P = 2.9×10^−4^)^18;30^.

**Figure 2:**
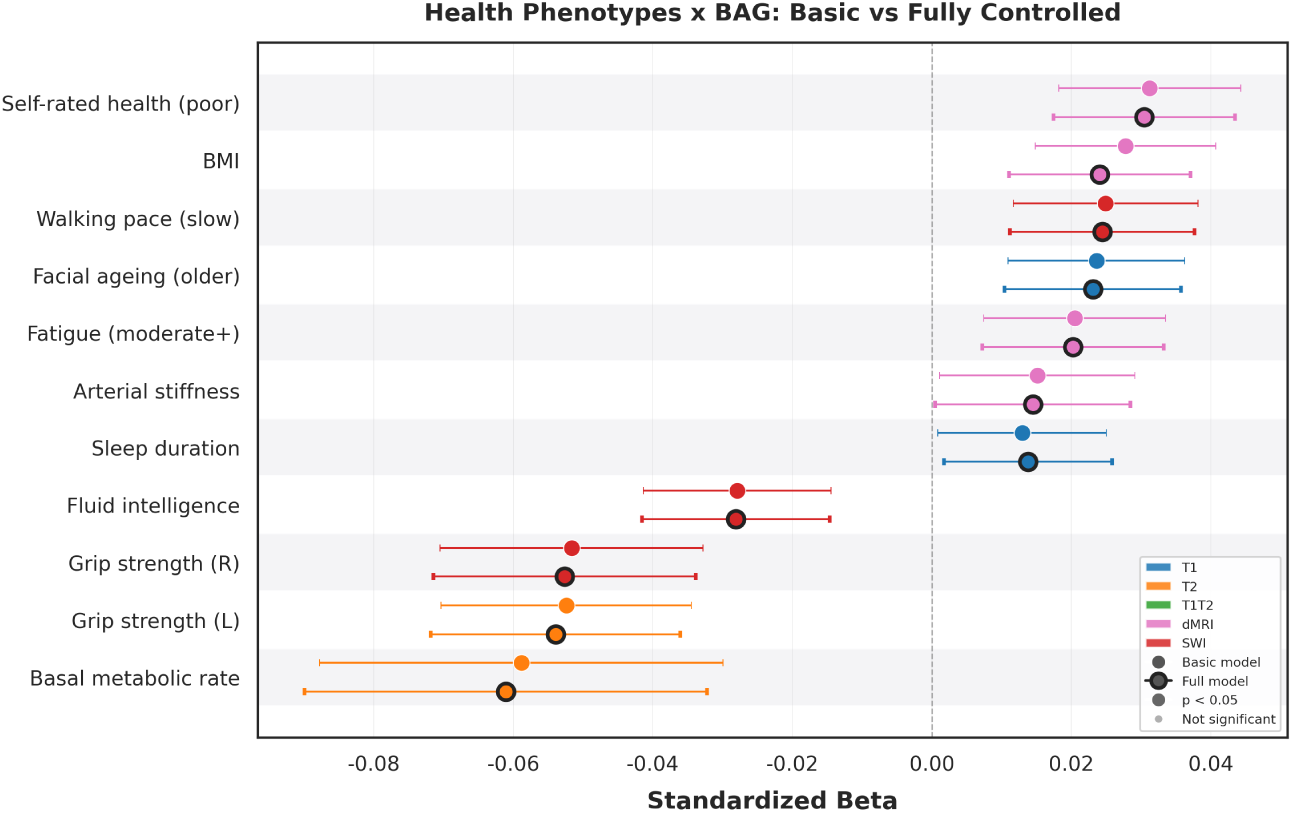
Health-phenotype associations of BAG. Points and 95% CIs are standardised *β* coefficients per s.d. increase in BAG, for basic (age + sex) and fully controlled (age + sex + ethnicity + assessment centre + BMI) models. Modality colour: T1 (blue), T2 FLAIR (orange), T1+T2 fusion (green), dMRI (pink), SWI (red). Bold markers indicate the fully controlled model; black-edged markers reach P < 0.05.

Modality-specific effects were biologically coherent. Arterial stiffness index (ASI), a non-invasive vascular-aging measure, was significantly associated only with dMRI BAG (*β* = +0.015, P = 0.04) — consistent with dMRI’s unique sensitivity to white-matter microstructure, which is vulnerable to chronic hypoperfusion and increased pulsatile energy transmission from stiffened arteries ^17;31^. Adding BMI, smoking status and alcohol frequency to the basic age-and-sex model changed *β* by < 10% across all modality–outcome pairs, indicating that BAG–health associations are not driven by these lifestyle confounds.

### Cognitive function analysis

Across 49 cognitive tests, SWI BAG consistently produced the largest standardised effect sizes (Fig. 3). The strongest single-test association was for reaction time, where SWI BAG was associated with longer reaction times (*β* = +0.058, P = 4.8×10^−15^) versus *β* = +0.045 for T1 (P = 3.9×10^−11^). For Symbol Digit Substitution — the test with the most consistent multimodal signal — modality *β* values for correct responses ranged from −0.052 (T1) to −0.060 (dMRI), all P < 10^−11^. Matrix pattern completion (SWI *β* = −0.052 vs. T1 *β* = −0.041) and Trail-Making A duration (SWI *β* = +0.061 vs. T1 *β* = +0.034) showed similar SWI > T1 ordering; for fluid intelligence (p20016) SWI BAG showed the largest effect (*β* = −0.026), ∼3× that of T1. The biological basis of this SWI amplification likely resides in SWI’s sensitivity to brain iron, which accumulates with age in basal ganglia, thalamus and substantia nigra, catalysing oxidative stress through Fenton chemistry and contributing to neuronal vulnerability ^32–34^. dMRI BAG showed intermediate cognitive effect sizes — stronger than T1 for fluid intelligence, symbol-digit and Trail-Making B — consistent with the role of white-matter microstructure in efficient neural transmission ^17;29^. Direction of association was concordant across all five modalities for 85.7% of cognitive tests (42 / 49), and at least four modalities reached P < 0.05 for 80% of tests; crystallised-knowledge measures (e.g., picture vocabulary) showed no significant BAG associations in any modality, consistent with cognitive-reserve theory ^18^.

**Figure 3:**
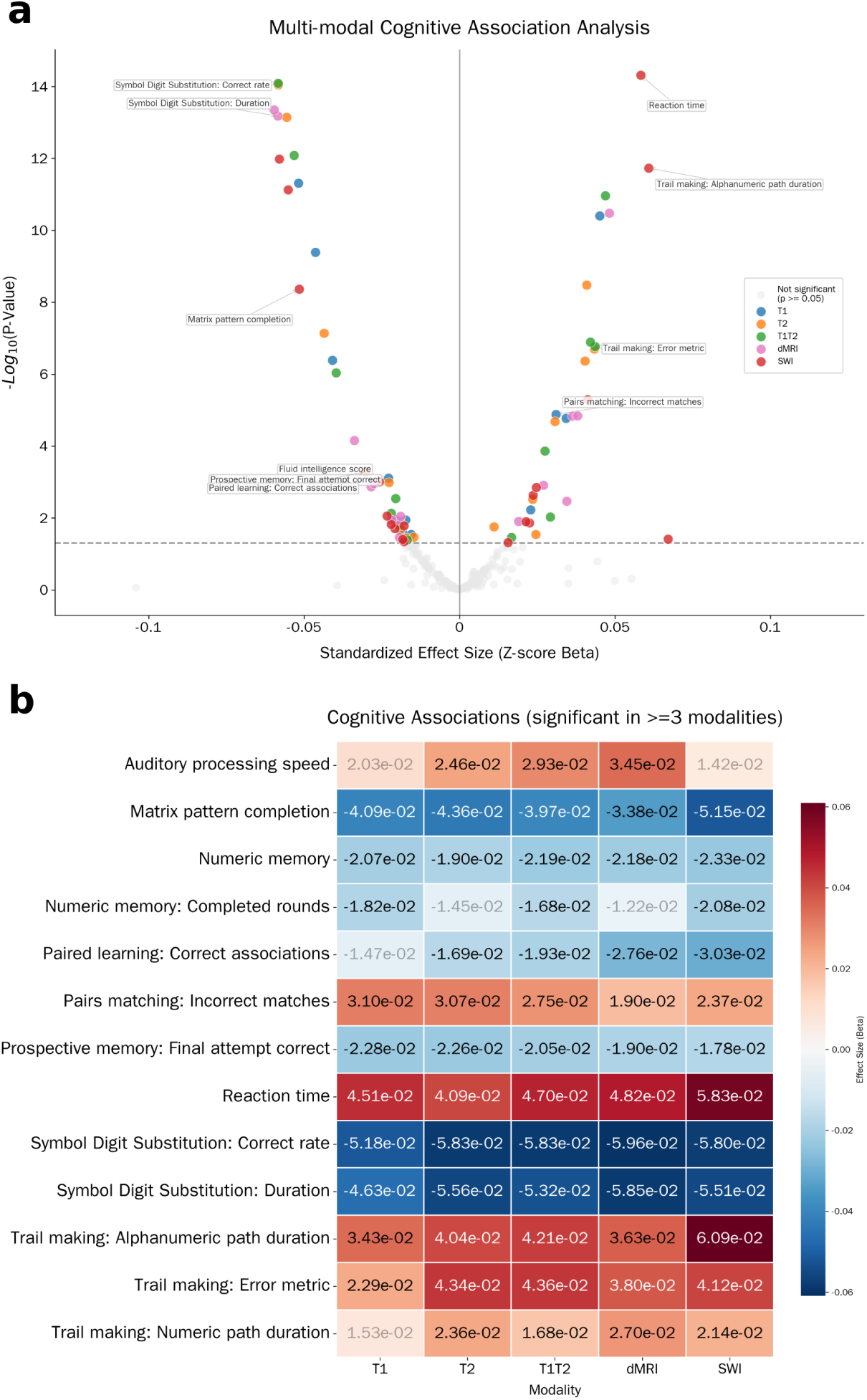
Cognitive associations across modalities. **a**, Volcano plot of effect size (standardised *β*) versus −log_10_(P) across 49 cognitive tests for the five modalities; top tests are annotated. **b**, Heat-map of standardised *β* for the 13 tests significant in three or more modalities (FDR q < 0.05); cell colour encodes *β* and cell text the value.

### Disease risk

To evaluate the prognostic value of modality-specific BAG, we performed Cox proportional hazards regression for 26 disease outcomes ascertained from linked hospital episode statistics, inpatient admissions and death-registry data using ICD-10 codes (Methods). For each outcome, the first recorded diagnosis after the imaging visit was used as the event time; participants were censored at last follow-up (median 5.2 yr). Models were stratified by sex and adjusted for chronological age at imaging and ethnicity; analyses were restricted to the 18,107 participants with data on all five modalities to enable fair cross-modality comparison. Hazard ratios (HR) are reported per one s.d. increase in BAG (Fig. 4).

**Figure 4:**
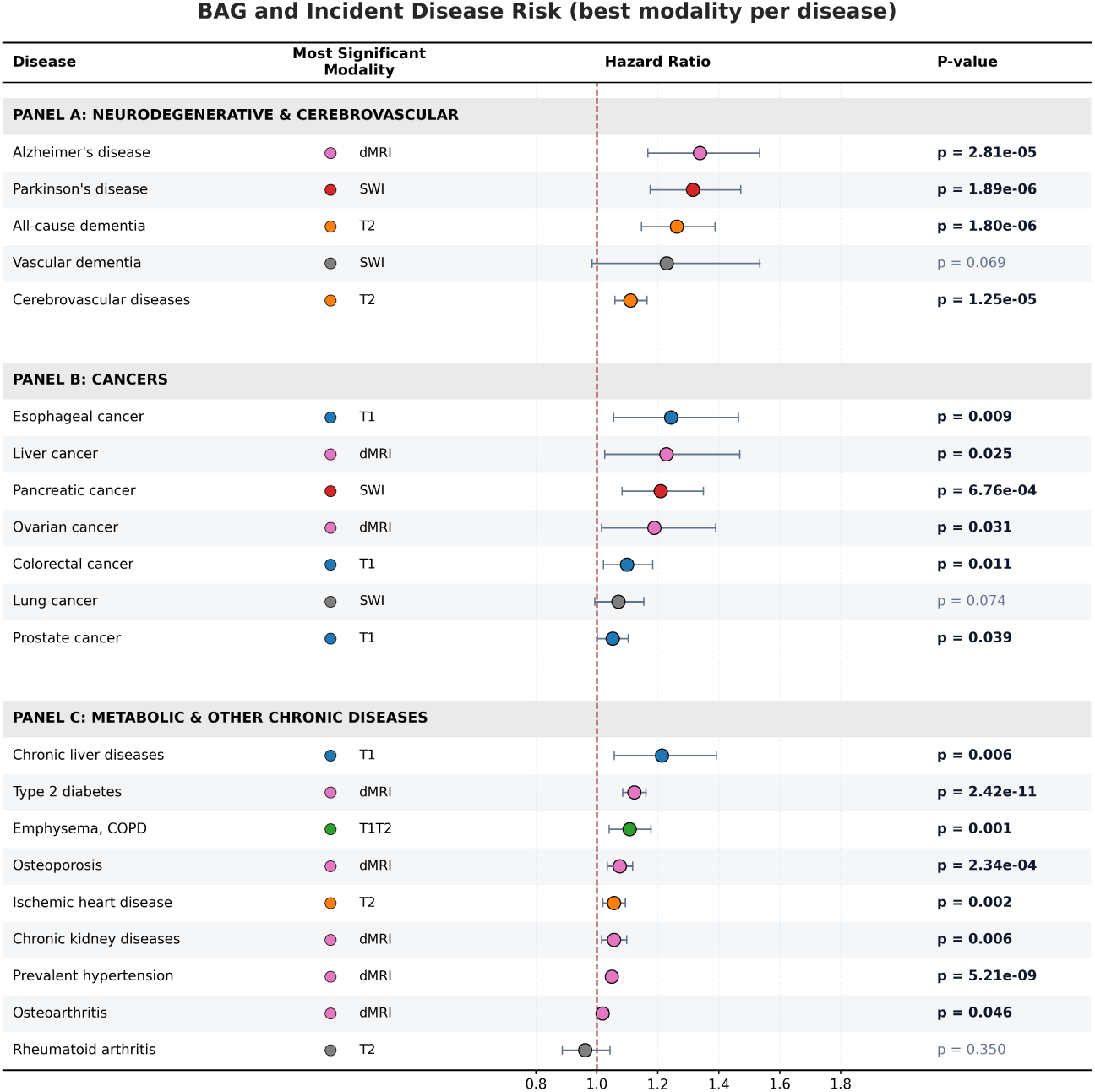
Forest plot of BAG–disease associations: most significant modality per disease (per s.d. BAG; Cox proportional hazards adjusted for age and ethnicity, stratified by sex; n = 18,107). Panels group neurodegenerative/cerebrovascular, cancer and metabolic/chronic outcomes. Coloured markers indicate the modality giving the smallest P-value; black-text P-values denote P < 0.05.

Among **neurodegenerative outcomes**, BAG in every modality predicted incident dementia, Alzheimer’s disease and Parkinson’s disease, with clear modality specificity. T2 FLAIR BAG was the strongest predictor of all-cause dementia (HR 1.26, 95% CI 1.15–1.39, P = 1.8×10^−6^; 61 events). For Alzheimer’s disease (21 events), dMRI BAG carried the largest hazard (HR 1.34, 95% CI 1.17–1.53, P = 2.8×10^−5^), with T2 FLAIR a close second (HR 1.31). For Parkinson’s disease (29 events), T1 BAG produced the largest point estimate (HR 1.39, 95% CI 1.21–1.60, P = 2.4×10^−6^) while SWI BAG gave the smallest P-value (HR 1.32, P = 1.9×10^−6^) — a dissociation that aligns with T1’s volumetric sensitivity to nigrostriatal atrophy and SWI’s sensitivity to substantia-nigra iron ^16;34^. **Cerebrovascular outcomes** preferentially tracked T2 FLAIR BAG (cerebrovascular disease HR 1.11, P = 1.3×10^−5^, 241 events; ischaemic heart disease HR 1.06, P = 1.9×10^−3^, 445 events), consistent with FLAIR’s direct visualisation of WMHs. **Metabolic and cardiometabolic outcomes** were led by dMRI BAG: type 2 diabetes (HR 1.12, P = 2.4×10^−11^; 302 events), prevalent hypertension (HR 1.05, P = 5.2×10^−9^; 1,361 events), chronic kidney disease (HR 1.06, P = 5.9×10^−3^) and osteoporosis (HR 1.08, P = 2.3×10^−4^). **Cancer outcomes**, taken together, were largely not associated with BAG: of 15 cancer types tested, only pancreatic cancer (SWI HR 1.21, P = 6.8×10^−4^; 29 events), colorectal (T1 HR 1.10, P = 0.011), prostate (T1 HR 1.05, P = 0.038), oesophageal (T1 HR 1.24, P = 9.2×10^−3^) and liver (dMRI HR 1.23, P = 0.025) reached nominal significance, none surviving FDR correction across the cancer panel. This selective absence argues against a generic-frailty interpretation of BAG and reduces concern that the neurodegenerative findings are driven by prodromal disease ^18^.

Cumulative-incidence curves stratified by BAG decile (top 10%, middle 80%, bottom 10%) confirmed the Cox-based findings (Fig. 5). For prevalent hypertension, 8-year cumulative incidence reached ∼17% in the high-BAG decile versus ∼7% in the low decile; for type 2 diabetes, ∼6% versus ∼2%; for all-cause dementia, ∼1.0% versus ∼0.2%; for Parkinson’s disease, ∼0.6% versus ∼0.1%. For 21 of 26 outcomes, BAG-decile stratification produced significant separation (log-rank P < 0.05), demonstrating that the prognostic signal in BAG is broad rather than restricted to a few outcomes. A participant in the top decile of T2 FLAIR BAG carried an ∼5-fold higher 8-year cumulative incidence of dementia compared with the bottom decile — a risk gradient that approaches the effect of a decade of chronological age — suggesting that multimodal BAG profiling could enrich Alzheimer’s prevention trials with the highest-risk individuals using routinely acquired MRI ^35;36^.

**Figure 5:**
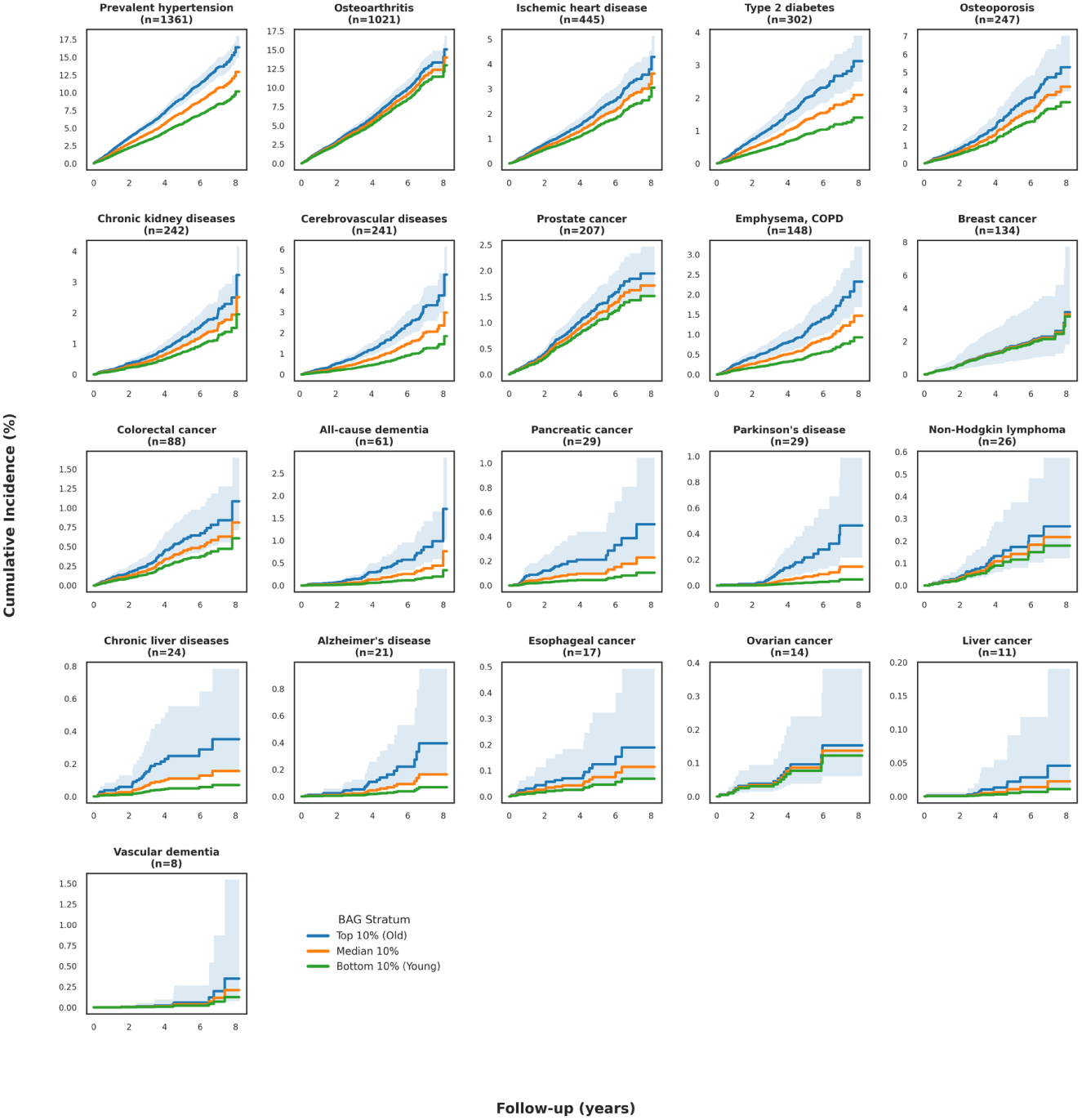
Cumulative-incidence curves stratified by BAG decile (top 10% blue; median 80% orange; bottom 10% green) for 21 disease outcomes spanning neurodegenerative, cerebrovascular, metabolic and cancer categories. Shaded bands denote 95% confidence intervals.

### Neuroanatomical interpretation

To identify the brain regions and tissue compartments driving age prediction across modalities, we generated Grad-CAM attribution maps from the final convolutional layer of DenseNet121 for held-out test participants across all five folds, quantifying attribution within three tissue classes (grey matter, white matter, CSF) defined by FSL FAST and within 14 subcortical regions defined by FSL FIRST.

For T1, Grad-CAM attributed 47.5% of total absolute attribution to grey matter, 25.6% to white matter and 26.9% to CSF; this pattern was qualitatively conserved across modalities, with the grey-matter share ranging from 44.6% (T1+T2 fusion) to 47.5% (T1, SWI) and the CSF share from 22.5% (T1+T2 fusion) to 27.4% (T2 FLAIR, dMRI; Fig. 6b). The substantial CSF attribution reflects Grad-CAM’s known sensitivity to high-contrast boundaries — particularly the CSF-pial and CSF-ependymal interfaces, where intensity gradients are maximal ^26;32^.

**Figure 6:**
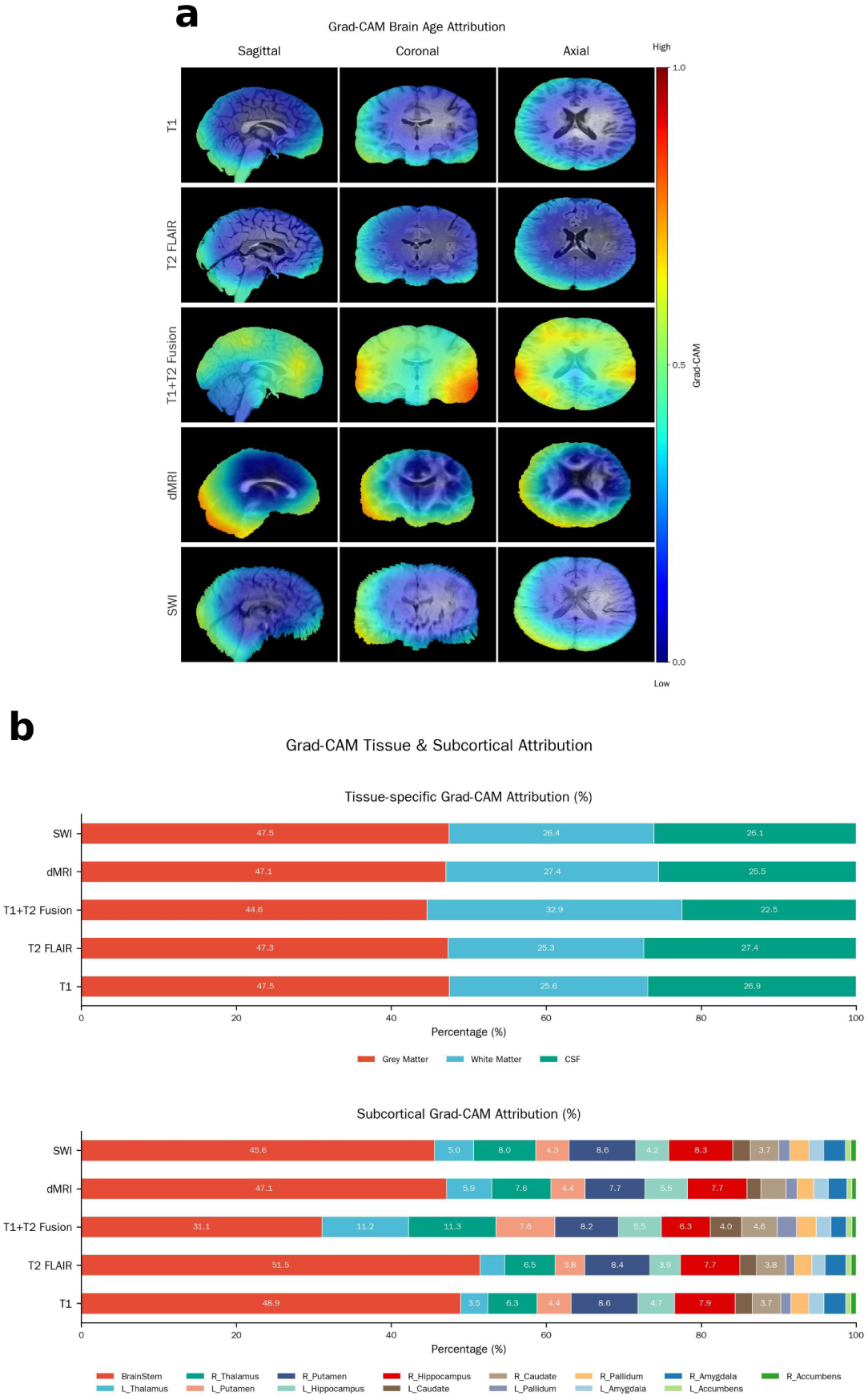
Neuroanatomical interpretation. **a**, Grad-CAM saliency maps overlaid on participant MRI for the five modalities (sagittal, coronal, axial). **b**, Tissue-class and subcortical Grad-CAM attribution per modality. Images reproduced by kind permission of UK Biobank ©.

The T1+T2 fusion model produced a notable redistribution: white matter rose from 25.6% under T1 alone to 32.9% under fusion — a relative increase of ∼29% — while grey matter fell from 47.5% to 44.6% and CSF from 26.9% to 22.5%. We interpret this as **representational synergy**: the joint T1+T2 input provides intensity signatures that more reliably distinguish white-matter pathology (visible on T2) from normal volumetric ageing (better characterised by T1 atrophy patterns), letting the model rely less on edge-based cortical features and more on tissue-internal signal characteristics. To our knowledge this is the first quantitative demonstration that multimodal fusion reshapes — rather than linearly combines — the spatial features driving brain-age prediction.

Subcortical attribution revealed a consistent hierarchy across modalities (Fig. 6b). The thalamus and putamen dominated (each ∼3–11% of total subcortical attribution), followed by hippocampus, caudate and pallidum. This hierarchy maps onto known age-related vulnerability: the thalamus shows pronounced age-related volume loss and iron accumulation; the putamen is a primary site of age-related iron deposition visible on SWI ^33;34^; and the hippocampus is the epicentre of Alzheimer’s pathology and a region of accelerating atrophy with advancing age ^11;37^. Disease–anatomy mapping reinforces biological plausibility: the putamen, high in attribution across all modalities, is the primary site of Parkinson’s degeneration, consistent with T1 BAG most strongly predicting Parkinson’s risk; the hippocampus, consistently weighted in T1 and T2 models, aligns with T2 FLAIR and dMRI BAG predictions of dementia; and the thalamus — the most-weighted subcortical region — is a hub of cerebral small-vessel disease, consistent with T2 FLAIR BAG’s strong prediction of cerebrovascular outcomes ^16;17^.

### Causal effects of lifestyle on brain age

Having established that BAG is a clinically meaningful and neuroanatomically grounded biomarker, we asked whether modifiable lifestyle behaviours causally accelerate or attenuate brain aging. We performed two-sample summary-data Mendelian randomization (MR) for seven lifestyle exposures: cooked vegetable intake, fresh fruit, oily fish, coffee intake, tea intake, moderate-to-vigorous physical activity (MVPA), alcohol intake frequency and smoking initiation (Methods). Independent genetic instruments (*P* < 5 × 10^−8^; LD-pruned at *r*^2^ < 0.001 within 10,000 kb windows) were drawn from UKB GWAS summary statistics, and SNP–outcome effects were computed for T1, T2 FLAIR and T1+T2 BAG. Causal estimates were obtained with three complementary estimators — inverse-variance-weighted (IVW; primary), MR-Egger and weighted-median — and pleiotropy was assessed via the MR-Egger intercept.

Genetically predicted physical activity exerted a robust protective causal effect on BAG across modalities (T1+T2 *β* = −0.29 yr per s.d. exposure, P = 1.1×10^−19^; Fig. 7a), as did oily fish intake (T1+T2 *β* = −0.20, P = 6.5×10^−20^), fresh fruit (T1+T2 *β* = −0.22, P = 9.2×10^−23^) and coffee intake (T1+T2 *β* = −0.16, P = 4.3×10^−14^).

**Figure 7:**
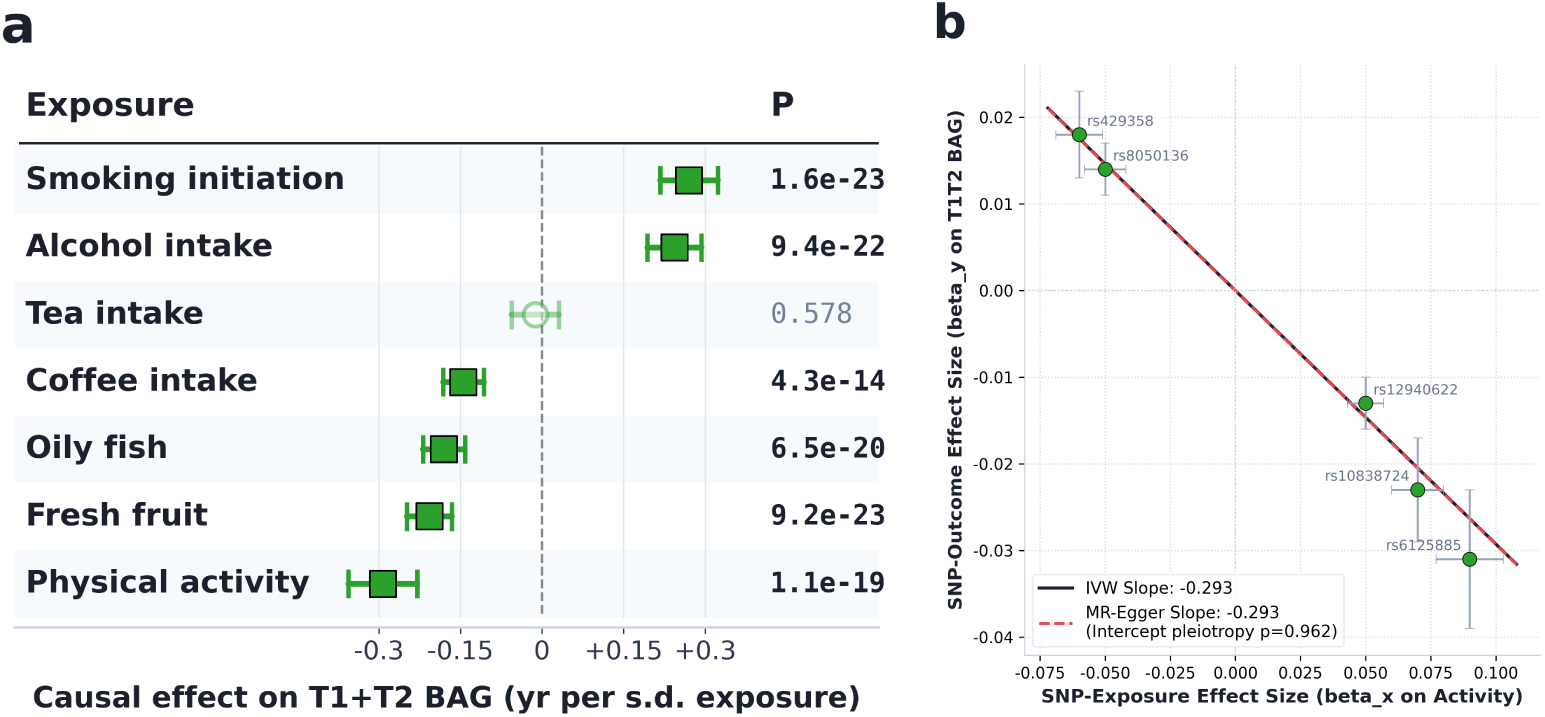
Causal effects of lifestyle on T1+T2 BAG from two-sample summary-data Mendelian randomization. **a**, Tabular forest plot of inverse-variance-weighted (IVW) causal estimates for seven lifestyle exposures on T1+T2 fusion BAG, expressed as years of BAG per s.d. increase in the genetically predicted exposure. Rows are ordered from most aging-accelerating (top) to most protective (bottom); filled squares with whiskers denote the point estimate and 95% confidence interval; open circles indicate non-significant effects (P ≥ 0.05). **b**, SNP-level scatter for the strongest protective exposure (physical activity) on T1+T2 BAG; IVW and MR-Egger slopes are essentially identical and the MR-Egger intercept P = 0.96, indicating no directional pleiotropy.

Conversely, smoking initiation and alcohol intake frequency causally accelerated brain aging (smoking T1+T2 *β* = +0.25, P = 1.6×10^−23^; alcohol T1+T2 *β* = +0.24, P = 9.4×10^−22^); tea intake showed no significant causal effect (P > 0.5 across estimators). IVW and MR-Egger slopes were virtually identical for the strongest exposures — for example, the physical-activity instrument yielded IVW slope −0.293 with MR-Egger slope −0.293 and intercept-test P = 0.96 (Fig. 7b) — and MR-Egger intercept tests were non-significant across all exposures, supporting the absence of directional horizontal pleiotropy and the validity of the instruments. These genetically anchored estimates provide convergent causal evidence that beneficial lifestyle behaviours attenuate, while smoking and alcohol accelerate, structural brain aging.

## Discussion

The central finding of this study is that brain age, as estimated by deep-learning models, is not a unitary construct: different MRI contrasts capture distinct, biologically meaningful facets of brain aging, each with a characteristic profile of health associations, cognitive correlates, prognostic value, neuroanatomical basis and causal lifestyle determinants. These findings challenge the prevailing assumption that brain age can be adequately summarised by a single metric from a single contrast.

Our study makes four core contributions. First, we provide what is, to our knowledge, the largest within-cohort head-to-head comparison of deep-learning brain-age models across five MRI modalities in up to 24,648 UKB participants, with external validation in PPMI. Prediction accuracy alone is an incomplete metric of a modality’s value: T1+T2 fusion achieved the highest accuracy (MAE 2.19 yr), yet dMRI BAG — with ∼28% higher prediction error — uniquely captured arterial stiffness and most strongly predicted incident type 2 diabetes (HR 1.12, P = 2.4×10^−11^), and SWI BAG, with the highest error of all five modalities, showed the largest cognitive effect sizes. The “best” brain-age model therefore depends on the intended use case: for dementia risk stratification, T2 FLAIR; for Parkinson’s disease, T1; for cognitive assessment, SWI; for vascular–metabolic profiling, dMRI. Second, through comprehensive phenotyping across 49 cognitive tests, 11 health measures and 26 disease endpoints we demonstrate that BAG is a trans-diagnostic biomarker with broad prognostic utility. The hierarchical organisation of disease risk — strongest for neurodegenerative conditions (HR 1.26–1.39), intermediate for cerebrovascular and metabolic disease (HR 1.05–1.12) and largely absent for cancer — indicates that BAG captures tissue-specific aging rather than a generic frailty signal ^4;18^. The convergence between SWI’s cognitive amplification and the SWI–pancreatic-cancer signal (HR 1.21, P = 6.8×10^−4^) is intriguing and may point to iron-related mechanisms — ferroptosis and oxidative stress — shared between neurodegeneration and certain cancers ^33^. Third, our Grad-CAM analyses show that the T1+T2 fusion model qualitatively reshapes — rather than linearly combines — the spatial features driving age prediction, shifting ∼7 percentage points of attribution toward white matter relative to T1 alone, a phenomenon we term *representational synergy*. Fourth, two-sample MR analyses provide genetically anchored evidence that protective behaviours — physical activity, oily fish, fresh fruit and coffee intake — causally attenuate, while smoking initiation and alcohol intake frequency causally accelerate, structural brain aging. By using independent genetic variants as instrumental variables we bypass residual confounding and reverse causation, identifying concrete, genetically validated targets for lifestyle and public-health interventions aimed at preserving cognitive reserve and slowing structural brain decline ^1;22^.

Several limitations should be considered. UKB is a volunteer cohort with healthy-volunteer bias ^27^, which may compress BAG variance and attenuate associations relative to more representative or clinical populations; replication in additional population-based and disease-enriched cohorts beyond PPMI remains an important next step. Our brain-age models are trained cross-sectionally rather than longitudinally; within-person change may yield a more sensitive measure of accelerated aging ^18^. While we provide evidence that SWI BAG captures an iron-related signal and dMRI BAG captures microstructural integrity, direct validation with quantitative imaging biomarkers — R2* relaxometry or quantitative susceptibility mapping for iron, and neurite-orientation dispersion or free-water imaging for diffusion — is needed to establish the specific biological correlates of each modality’s BAG ^16^. Finally, our MR analyses use UKB itself for both exposure and outcome instruments, raising the possibility of sample overlap that can bias two-sample MR toward the observational estimate; replication using non-UKB GWAS for the exposures is an important next step ^23^.

These limitations notwithstanding, our findings carry direct clinical and methodological implications. Clinically, multimodal BAG profiling could contribute to risk stratification in the growing landscape of Alzheimer’s prevention trials ^35;36^, identifying individuals with accelerated brain aging up to a decade before clinical onset using routinely acquired MRI; the modality specificity of associations suggests that different preventive strategies may be indicated depending on which contrast carries the elevated BAG. Methodologically, our results argue that the field should move beyond single-modality, single-metric brain age toward multi-contrast frameworks that acknowledge the multi-faceted nature of brain aging, and pair model performance with attribution maps that quantify which compartments drive the prediction. The causal lifestyle estimates further argue that brain-age biomarkers are not merely diagnostic mirrors but actionable readouts: physical activity, healthy diet, moderation of alcohol and avoidance of smoking measurably and causally slow structural brain aging.

## Methods

### Participants and imaging data

We used data from the UK Biobank imaging enhancement, acquired between 2014 and 2023 on four identical Siemens Skyra 3T scanners under a harmonised protocol ^16;19;27^. Brain MRI sequences included T1-weighted MPRAGE (1 mm isotropic; TE/TR = 2.01/2000 ms; TI = 880 ms; flip angle 8°), T2 FLAIR (1.05 × 1 × 1 mm; TE/TR = 399/5000 ms; TI = 1800 ms), dMRI (2 mm isotropic; b = 1000 and 2000 s/mm^2^; 100 directions) and SWI (0.8 mm isotropic; TE/TR = 20/29 ms; flip angle 15°). All images were preprocessed with the UKB brain-imaging pipeline (v2) including gradient-distortion correction, rigid-body motion correction and brain extraction^19^. For our models, images were further registered to MNI152 1 mm standard space using affine transformation only (FSL FLIRT, 12 d.f., normalised mutual information cost function) — chosen because nonlinear registration was empirically shown to attenuate age-related morphology (see Results). Use of UKB data was approved under application 89483. UK Biobank obtained informed consent from all participants, and the present analyses used de-identified data under UK Biobank access procedures.

### Model architecture and training

We used a 3D DenseNet121 adapted for single-channel (T1, T2, dMRI, SWI) or dual-channel (T1+T2 fusion) input, comprising an initial convolution and pooling, four dense blocks (channels 64, 128, 256, 512) connected by transition layers, global average pooling and a linear output. Implementation used MONAI 1.3 / PyTorch 2.1 with mixed-precision training. Training used five-fold cross-validation stratified by age decile, MAE loss, the Adam optimiser (1 × 10^−4^ learning rate; cosine annealing), batch size 96 and distributed data parallel across three NVIDIA RTX 4090 GPUs. Models were trained for 240 epochs (single modalities) or 490 epochs (T1+T2 fusion); the epoch with the lowest validation MAE was retained.

### External validation (PPMI)

External validation used the Parkinson’s Progression Markers Initiative (PPMI) for T1 and T2 FLAIR. PPMI was preprocessed with the same affine-only registration pipeline used for UKB (`fslreorient2std` → `robustfov` → BET (-f 0.3 -R) → FLIRT (6 d.o.f. for masking, 12 d.o.f. for normalisation) → `fslmaths -mas`). PPMI Philips scans with non-unit `PhilipsRescaleSlope` were rescaled prior to registration. External validation was confined to T1 and T2 FLAIR because the dMRI and SWI sequences in PPMI are not directly compatible with the UKB acquisition protocol.

### Statistical analysis

BAG was defined per modality as predicted age minus chronological age. Bias correction for regression-to-the-mean was applied using the method of Liang et al. ^28^: BAG_corrected_ = BAG_raw_/(1 − *r*^2^) where *r* is the within-modality prediction correlation. Phenotypic associations used ordinary least squares with robust (Huber–White) standard errors, adjusted for age, sex, ethnicity, assessment centre and BMI. Effect sizes are standardised *β* coefficients. Cox proportional hazards regression used age at imaging as the time scale with Efron’s approximation for tied events, stratified by sex and adjusted for ethnicity. The proportional-hazards assumption was verified using Schoenfeld residuals (P > 0.05 for all reported models). Multiple comparisons were controlled with the Benjamini–Hochberg FDR procedure (q < 0.05) within each analysis family (cognitive tests, health phenotypes, disease outcomes).

### Two-sample Mendelian randomization

Independent genetic instruments associated with each exposure at genome-wide significance (*P* < 5 × 10^−8^) and low linkage disequilibrium (*r*^2^ < 0.001 within 10,000 kb windows) were selected from UKB GWAS summary statistics for cooked vegetables, fresh fruit, oily fish, coffee intake, tea intake, MVPA, alcohol intake frequency and smoking initiation. SNP–outcome effect sizes (*β*_*Y*_, *se*_*Y*_) were obtained for T1, T2 FLAIR and T1+T2 BAG. The primary estimator was the inverse-variance-weighted (IVW) random-effects model:

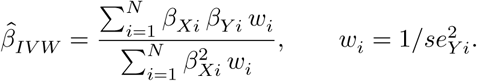

We additionally implemented MR-Egger regression to test directional horizontal pleiotropy by including an intercept *θ*_0_: *β*_*Y i*_ = *θ*_0_ + *β*_Egger_ *β*_*Xi*_ + *ɛ*_*i*_, with *θ*_0_ ≠ 0 (*P* < 0.05) indicating pleiotropy. We also used the weighted-median estimator, which is consistent when < 50% of weight comes from invalid instruments; its standard error was estimated by 500 bootstrap iterations. All MR analyses were implemented in a custom Python pipeline using GWAS summary-level data only.

### Attribution mapping

Grad-CAM was computed using the MONAI implementation with the final convolutional layer of DenseNet121 as the target^26^. Per-voxel attributions were averaged across all test-set participants within each cross-validation fold and then across folds. Region-wise attribution was quantified by summing absolute attribution within each anatomical region defined by FSL FAST (tissue segmentation) and FSL FIRST / Harvard-Oxford probabilistic atlas (subcortical structures), divided by region volume to obtain mean absolute attribution density.

## Data availability

UKB data are available to bona fide researchers via application at https://www.ukbiobank.ac.uk. GWAS summary statistics used for MR are publicly available from the Neale Lab UKB release. Source data underlying every figure in this paper (CSV/Parquet) are provided in the project repository.

## Code availability

The infrastructure code (data loading, preprocessing, and UK Biobank phenotype utilities) is available at https://github.com/HumbleHumbert/ukbeaver. All analysis code — training pipelines, statistical analyses, MR pipelines and plotting scripts — is available at https://github.com/HumbleHumbert/ukbag, along with the source data files used to generate each figure.

## Author contributions

Ruize Yu: Methodology, Software, Visualization, Writing – original draft. Shanshan Shao: Conceptualization, Supervision, Writing – review & editing. Feng Xu: Data curation, Formal analysis, Validation, Project administration, Writing – review & editing.

## Acknowledgments

We thank Professor Steven Su (Rehabilitation Robotics Laboratory, Shandong First Medical University) for providing the computational resources used throughout this work. This research has been conducted using the UK Biobank Resource under Application Number 89483.

